# A Multilayer Model for Early Detection of COVID-19

**DOI:** 10.1101/2021.02.25.21252470

**Authors:** Erez Shmueli, Ronen Mansuri, Matan Porcilan, Tamar Amir, Lior Yosha, Matan Yechezkel, Tal Patalon, Sharon Handelman-Gotlib, Sivan Gazit, Dan Yamin

## Abstract

Current efforts for COVID-19 screening mainly rely on reported symptoms and potential exposure to infected individuals. Here, we developed a machine-learning model for COVID-19 detection that utilizes four layers of information: 1) sociodemographic characteristics of the tested individual, 2) spatiotemporal patterns of the disease observed near the testing episode, 3) medical condition and general health consumption of the tested individual over the past five years, and 4) information reported by the tested individual during the testing episode. We evaluated our model on 140,682 members of Maccabi Health Services, tested for COVID-19 at least once between February and October 2020. These individuals had 264,516 COVID-19 PCR-tests, out of which 16,512 were found positive. Our multilayer model obtained an area under the curve (AUC) of 81.6% when tested over all individuals, and of 72.8% when tested over individuals who did not report any symptom. Furthermore, considering only information collected before the testing episode – that is, before the individual may had the chance to report on any symptom – our model could reach a considerably high AUC of 79.5%. Namely, most of the value contributed by the testing episode can be gained by earlier information. Our ability to predict early the outcomes of COVID-19 tests is pivotal for breaking transmission chains, and can be utilized for a more efficient testing policy.

## 1 Introduction

Severe acute respiratory syndrome coronavirus 2 (SARS-CoV-2 or COVID-19) was first identified in Wuhan, China, in December 2019. It has since developed into a pandemic, affecting 219 countries and territories worldwide, causing over 109 million cases and claiming over 2.4 million lives as of February 18, 2021^1^.

Despite the considerably fast development of an effective vaccine, the pandemic is expected to continue disrupt our lives in the near future for multiple reasons. These include the emergence of highly transmissible mutant strains^2,3^, the incomplete efficacy of the developed vaccines and their disapproval for use in certain populations^4^, the limited supply and distribution capacities of the vaccines^5^, as well as the potential risk of vaccine-waning immunity^6^. Thus, in parallel with the challenge of increasing vaccination coverage and long-term effectiveness, various efforts for early detection and prompt isolation are required to breaking transmission chains and containing local outbreaks.

Current efforts for early detection of COVID-19 mainly rely on screening practices, which typically include a combination of reported symptoms and potential exposure to infected individuals^7^. Among other symptoms, loss of taste and smell, fatigue, and fever may be present in COVID-19 patients and were found to be useful for detection^7,8^. However, provided that multiple pathogens may cause similar symptoms as COVID-19, symptoms-based detection is limited. Moreover, it is inherently prone to miss presymptomatic or asymptomatic cases, which account for 40–45% of those infected with COVID-19, and can still transmit the disease^7,9^. Consequently, the US has recently scaled up its efforts to improve testing capacity and accuracy in an unprecedented manner^10^.

Several pioneering studies have offered proactive methods for COVID-19 detection based on smartwatches, and activity trackers^11^–^13^. For example, a recent study showed that the integration of self-reported symptoms and sensor data from smartwatches resulted in an area under the curve (AUC) that was as high as 80%^11^. However, these methods rely on dedicated devices and require individuals’ cooperation in frequently wearing these devices and consenting to share the collected information. Such devices are used by less than 20% of the population in developed countries, and are also limited to specific age groups and subpopulations. Thus, it is crucial to improve our ability to detect the disease using already available data for the entire population.

As the risk of becoming infected is governed by the individuals’ contact mixing patterns, it is crucial to account for the disease’s spatiotemporal dynamics as part of the detection task^14,15^. Furthermore, certain populations are known to have a greater risk than others to test positive. Specifically, beyond age and gender, of great concern are the data showing the disproportionate effect of COVID-19 on ethnic and racial minorities, and impoverished populations^16,17^. These populations often live in denser regions are characterized by larger household sizes; thereby, they are at elevated risk of becoming infected^18^. The risk of contracting the disease also depends on an individual’s protective behavior, such as maintaining social distancing and improving hygiene practices. The latter correlates with the actual and perceived risks of an individual^19,20^, and both can be inferred from the individual’s medical history. Such evidence was also demonstrated in other contexts. For example, a previous study suggested that individuals who had not been vaccinated against influenza and were diagnosed with respiratory illness in the last season were more likely to become vaccinated in the upcoming season^21^. Under the same logic, information gained from the individual’s medical history that can be linked to the actual and perceived risks may serve as predictors for that individual’s test results.

Here, we developed a multilayer model for the early detection of COVID-19 infection. Our approach combines sociodemographic information about the tested individual, aggregated information on the spatiotemporal dynamics of the disease, and general information from the medical history of the individual, in addition to data collected during the testing episode. Our approach is pivotal for breaking transmission chains, and can be utilized to substantially improve testing strategies.

## 2 Results

Our study included a random sample of 140,682 individuals who were members of Maccabi Healthcare Services (MHS) and were tested for COVID-19 at least once between February and October 2020. Among these individuals, 53.8% were women, and their age ranged from 1 to 105 years, with a median age of 30 years (IQR: 16–49). These individuals had 264,516 COVID-19 tests, 16,512 (6.2%) of which were found to be positive.

Overall, we identified four layers of information that can help in predicting the outcome of a COVID-19 test: 1) the sociodemographic information of the tested individual, 2) the spatiotemporal patterns of the disease observed near the testing episode, 3) the medical condition and general health consumption of the tested individual over the past five years, and 4) the information collected from the tested individual during the testing episode. In examining the sociodemographic information of the tested individuals (Fig. 1A), we found that men were more likely to test positive than women, with 7.72±0.15% positive tests for men, compared to 5.11±0.11% for women. Positive tests were also linked with ethnicity and the socioeconomic level. Jewish orthodox and Arab individuals, who are characterized by larger household sizes, exhibited higher percentages of positive tests (14.2±0.35% and 7.78±0.4%, respectively) than the general population (4.66±0.09%). Individuals with low socioeconomic level presented a substantially higher percentage of positive tests (11.15±0.31%) than those with a medium or high levels (5.97±0.12% and 3.92±0.15%, respectively). Considering a predictive model based on this layer of information alone allowed a moderate classification ability between positive and negative tests, with an AUC of 67.74±0.77% (Fig. 3A).

**Figure 1.**
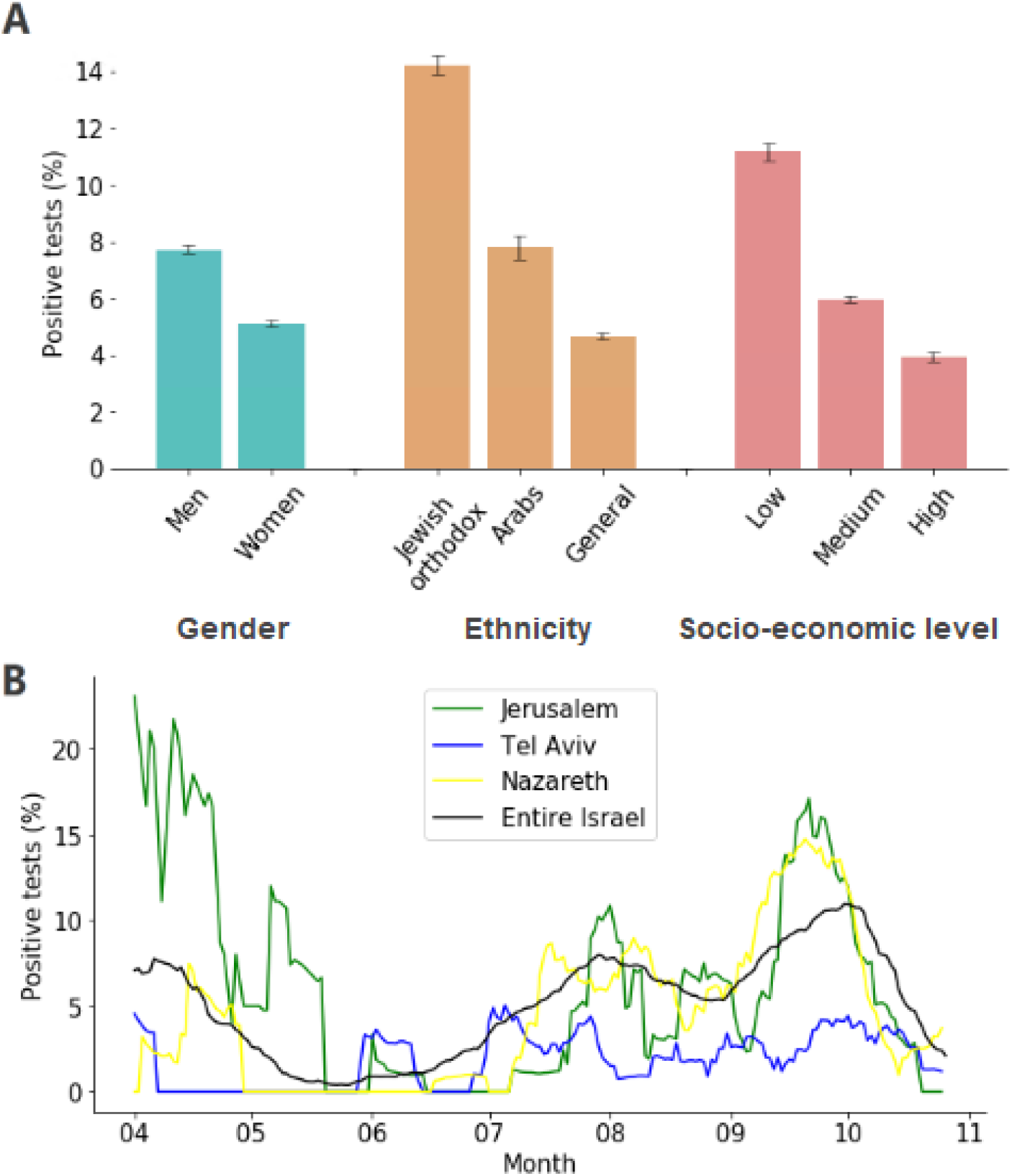
Layers 1 and 2 - sociodemographic information of the tested individual and spatiotemporal dynamics of the disease. (A) Percentage of positive tests stratified by gender, ethnicity, and socioeconomic level. The percentages of positive tests are linked with gender, ethnicity, and socioeconomic level. Error bars represent the 95% confidence interval. (B) Percentage of positive tests over time for three clinics located in different cities and for the entire country.

**Figure 3.**
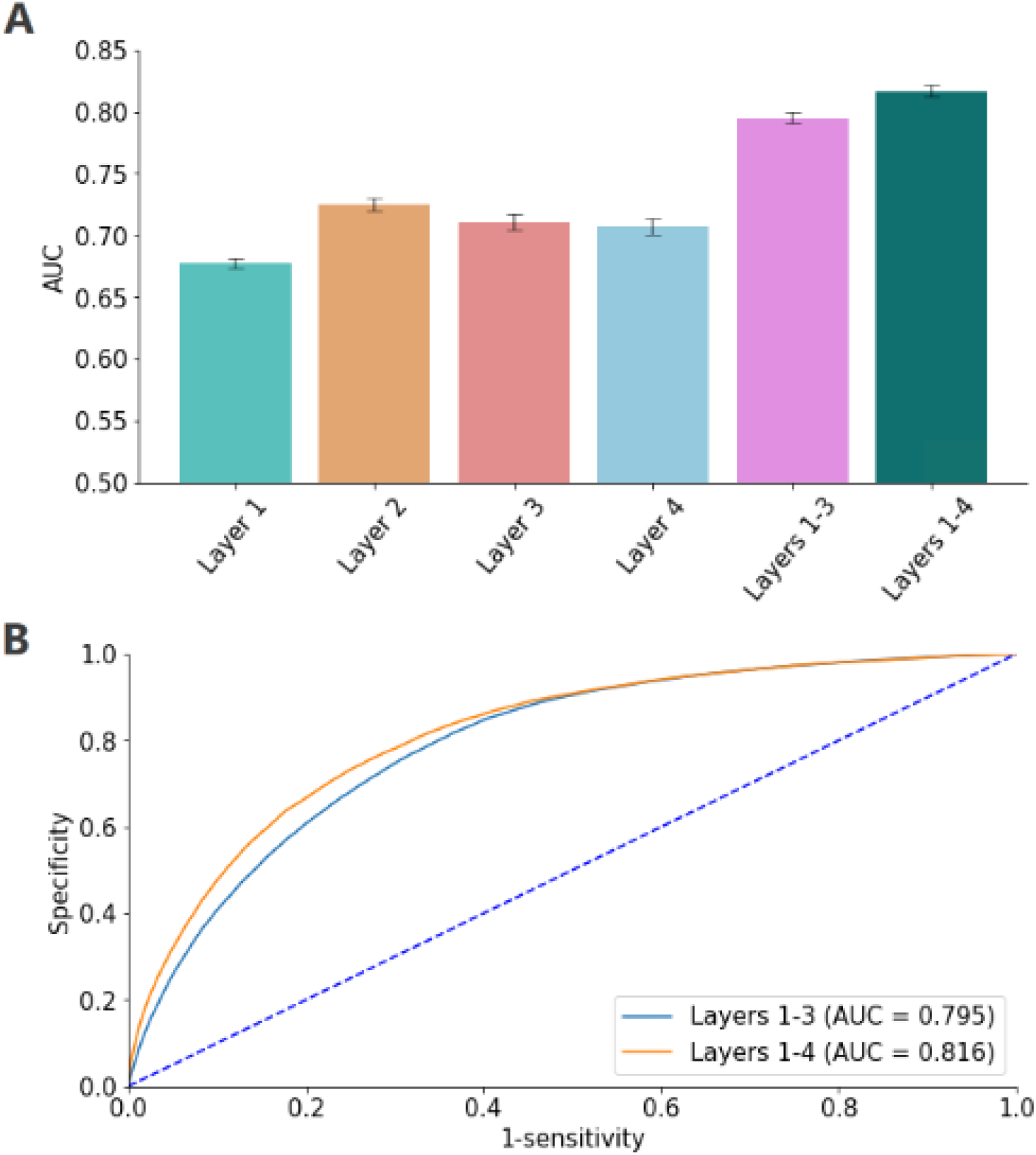
Predictive models’ performance. (A) Mean AUC of models based on layers 1-4 (sociodemographic information of the tested individual, spatiotemporal patterns of the disease, medical condition and general health consumption of the tested individual, and information collected during the testing episode) and the full model that combines all four layers. The full model allowed a considerably better classification ability between COVID-19-positive and COVID-19-negative tests with a mean AUC of 81.6%. Error bars represent the standard deviation of the 10 executions of the model. (B) Receiver operating characteristic curves for the full model and the model considering layers 1-3. The full model’s classification ability is only slightly better than that of the model considering the first three layers (i.e., excluding layer 4 - information collected during the testing episode).

The percentages of positive tests also varied considerably with time and across regions (Fig. 1B). Low percentages of positive tests characterized Tel Aviv compared to Jerusalem throughout most of the study period. Moreover, accounting for changes in time and region, we could identify regional outbreaks that were pivotal in our prediction task. For example, in specific zones in Nazareth, we observed lower rates than average in April but higher rates in October. Considering a predictive model based on this layer of information alone allowed a better classification ability between COVID-19-positive and COVID-19-negative tests with an AUC of 72.3± 0.44% (Fig. 3A).

Analyzing individuals’ electronic medical records (EMRs), we found that increased health consumption, increased preventative health behaviors, and particular medical conditions were associated with lower percentages of positive tests (Table 1). For example, individuals who were more likely to become vaccinated against influenza had a lower probability of testing positive across all age groups. For individuals aged 30-39, those who were vaccinated at least once over the past five years, were found positive in 4.34±0.35% of the tests, whereas those who were never vaccinated had 6.2±0.31% positive tests. Likewise, individuals who were diagnosed with cancer in the past had a lower probability of testing positive, across all age groups. Considering a predictive model based on this layer of information alone allowed a classification ability between positive and negative tests, with an AUC of 71±0.53% (Fig. 3A).

**Table 1.**
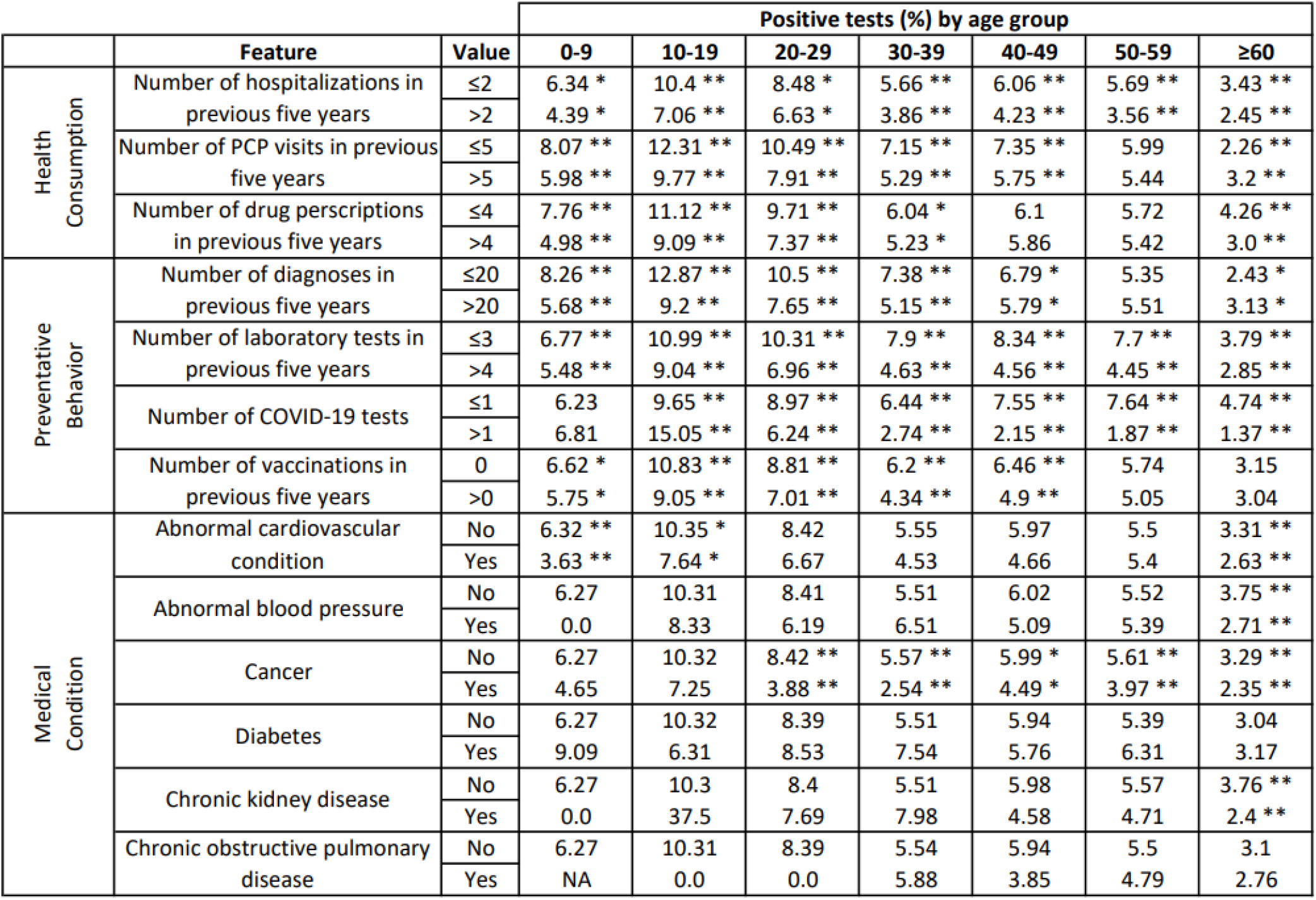
Layer 3 - health consumption, preventative health behavior and medical conditions. Percentages of positive tests stratified by health feature and age group. Significant differences are marked with stars, where ** denotes p<0.01 and * denotes p<0.05. Increased health consumption, increased preventative health behavior, and particular medical conditions, are associated with lower percentages of positive tests.

We also analyzed the information collected right before the COVID-19 test was taken, during the referral and during the testing episode itself. Specifically, we analyzed the association between reported symptoms and the test outcome (Fig. 2A). We found that loss of taste or smell was the most indicative symptom ranging from 10.52±0.05% of positive tests in individuals aged 0-9 to 33.16±0.03% in individuals aged 20-29. Interestingly, we found that several symptoms that are known to be caused by COVID-19 were negatively associated with a positive outcome. For example, the appearance of fever in children aged 0-9 may suggest a lower risk of testing positive, as there are many other causes of fever in this age group. Likewise, the existence of diarrhea in children aged 0-9 is less likely to be caused by COVID-19. We also found that exposure to infected individuals could serve as a predictor for the test outcome. Specifically, exposure to an infected individual at the same household was associated with an 18.48±0.64% chance of being found positive, and an exposure to other infected individuals (i.e., not in the same household) was associated with an 11.45±0.39% chance of being found positive (Fig. 2B). Moreover, we found that individuals who were tested at home had an elevated risk of being found positive (Fig. 2B). This is likely because testing at home was performed for individuals who were in quarantine or who suffered from a severe medical condition. Considering a predictive model based on this layer of information alone allowed a classification ability between positive and negative tests, with an AUC of 70.6±0.59% (Fig. 3A).

**Figure 2.**
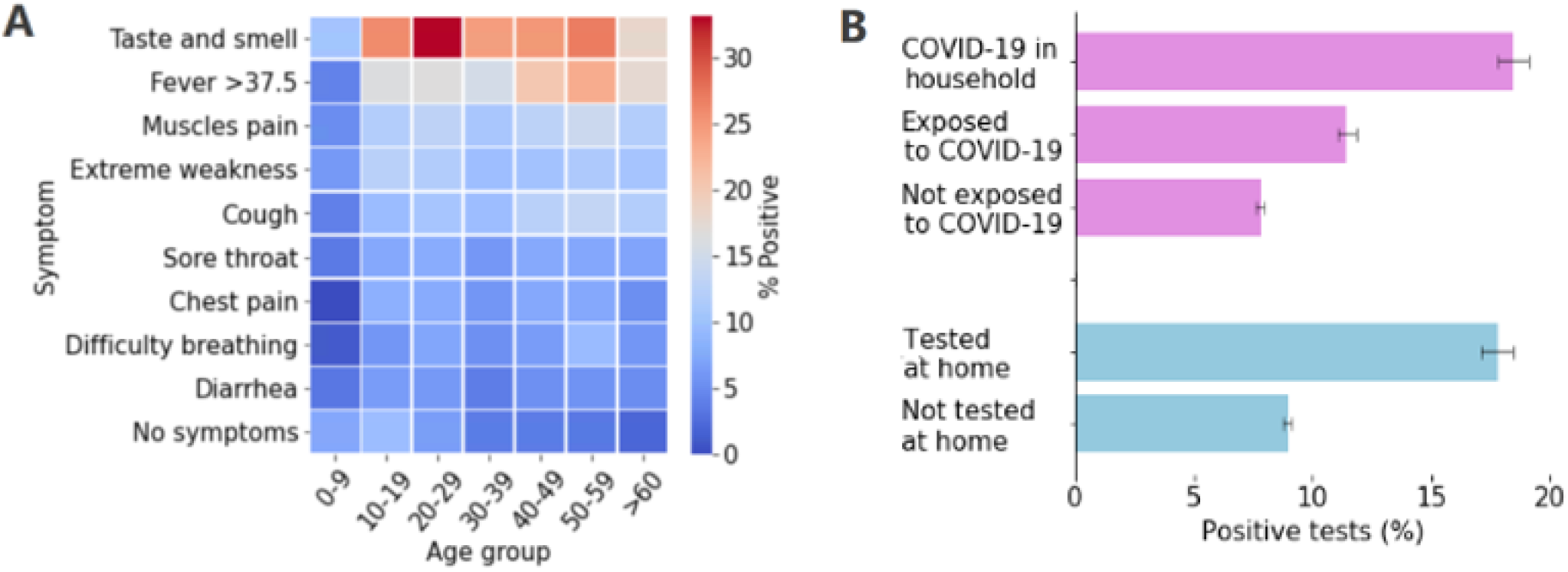
Layer 4 - information collected during the testing episode. (A) Percentages of positive tests stratified by symptoms and age groups. Several symptoms that are known to be caused by COVID-19 are negatively associated with a positive outcome. (B) Percentages of positive tests based on exposure to infected individuals, and on the test’s location. Individuals who were exposed to infected individuals and those who were tested at home had an elevated risk of being found positive.

Considering a predictive model that combines all four layers of information together allowed a considerably better classification ability between COVID-19-positive and COVID-19-negative tests, with an AUC of 81.6±0.46% (Fig. 3A). Notably, the classification ability of the full model was only slightly better than that of the model considering only information that was available prior to the testing episode (i.e., based on layers 1, 2, and 3), which yielded an AUC of 79.5± 0.6% (Fig. 3A). This marginal difference in performance between these two models can also be observed in Fig. 3B, which presents their full ROC curves. Lastly, limiting our predictions only to individuals who did not report any symptom, our full model yielded an AUC of 72.8±0.85%. This finding demonstrates a moderate, yet considerable, ability to identify individuals in their presymptomatic or asymptomatic clinical condition.

## 3 Discussion

We found that by using multiple layers of information, the risk of testing positive for COVID-19 is highly predictable, with the AUC reaching 81.6%. Specifically, we identified four layers of information that can predict positive COVID-19 test outcomes: 1) the sociodemographic characteristics of the tested individual, 2) the spatiotemporal patterns of the disease observed near the testing episode, 3) the medical condition and general health consumption of the tested individual over the past five years, and 4) the information reported by the tested individual during the testing episode.

We found that by analyzing information from the testing episode alone (e.g., symptom-related questions), we could achieve an AUC of 70.6%. This result is consistent (albeit lower) with recent studies that showed AUCs of 72%^11^ and 76%^7^. When we considered only the information collected before the testing episode – that is, before the individual may had the chance to report on any symptom – our model could reach a considerably higher AUC of 79.5%. This finding is pivotal for earlier detection. The marginal difference in AUC scores between the full model and the model without the testing episode information suggests that most of the information gained from the testing episode can be inferred by the individual’s medical history, as well as other aggregated information with regard to the disease dynamics. Moreover, while symptom-based predictions are likely to be sensitive to COVID-19 variants and the emergence of other respiratory infections, our approach is likely to be more robust, as it explicitly considers the spatiotemporal dynamics of COVID-19.

We found that individuals with underlying medical conditions and individuals who maintain a more preventative lifestyle are at lower risk of testing positive for COVID-19. This finding implies that those populations tend to protect themselves better against the disease or are more likely to be tested. While we cannot disentangle between the two causes, health behavior models, including the Health Belief Model^22^, and social cognitive theory^23,24^ suggest that the combination of these causes is likely. Despite the inherent differences in risk perceptions between cultures worldwide, we believe that the behavioral patterns and the predictive models we have developed can be reproduced with minor adaptations in most developed countries.

For privacy purposes, we considered only general information from EMRs to infer the individual’s health condition and preventative behavior. For example, in our model, we included information about the total number of yearly visits to PCP and the number of medications prescribed to a patient, rather than more invasive information such as the type of prescribed medication. Clearly, more detailed information about individuals may provide an improved understanding of their behavior and lead to improved predictive models. However, this comes with the price of invading privacy, which is not less important^25^.

Our study identifies and relies on correlations and associations in both pattern analysis and predictive modeling and does not attempt to assume or imply causality. In addition, this study does not explicitly account for the intervention efforts made by MHS during the study period, including efforts made to test individuals at higher risk. Third, the sensitivity and specificity of RT-PCR testing varies considerably in different age groups and considering the severity of the infection and the disease progression in the host^26^. Specifically, the sensitivity in mild cases could be as low as 62.5%^7,27^, and the sensitivity a day prior to symptom onset falls below 33%.

In conclusion, COVID-19 test results are highly predictable and can be achieved even in the absence of detailed information on the signs and symptoms of the individual during the testing episode. The ability to predict the outcomes of COVID-19 tests in real-time can be utilized for a more efficient testing policy. In the post vaccine era such a policy may become even more efficient due to lower transmission rates, enabling easier differentiation between positive and negative COVID-19 tests.

## 4 Methods

### 4.1 Ethical considerations

The study was approved by MHS’ Helsinki institutional review board, protocol number 0093-20-MHS, signed on October 21, 2020. Informed consent was waived as identifying details were removed before the analysis.

### 4.2 Study population and case definition

We analyzed the anonymized EMRs of 140,682 randomly sampled individuals tested at least once with PCR for COVID-19 during February-October 2020. The individuals were members of MHS. MHS is the second largest health maintenance organization (HMO) in Israel, serving more than 25% of the Israeli population (2.5 million members). MHS members are representative of the Israeli population and reflect all demographic, ethnic, and socioeconomic groups and levels^28^.

For the 140,682 individuals considered in this study, 279,140 COVID-19 tests were performed during the examined time period. According to previous guidelines in Israel, individuals who tested positive were motivated to conduct additional tests to terminate self-quarantine. Since our goal was to predict the presence of COVID-19, for each individual, we included in our analysis only tests until his/her first positive test (if such existed), which corresponded to 264,517 tests in total.

For each individual, we extracted data from their EMRs between 2015 and 2020. Specifically, we compiled four layers of information to predict COVID-19 test outcomes: 1) the sociodemographic information of the tested individual, 2) the spatiotemporal patterns of the disease, 3) the medical condition and general health consumption behavior of the tested individual, and 4) the information collected from the tested individual during the test procedure. Information on features considered for each of the layers is detailed in the Supplementary Information.

### 4.3 Statistical analysis

The problem of determining the outcome of a COVID-19 test (i.e., positive or negative) was treated as a machine learning, binary classification task. Specifically, we generated six different prediction models, based on single layers of information (sociodemographic, spatiotemporal, health-related and test-related), as well as on combination of layers (before the test, and before&during the test). We used XGBoost^29^ as the classification algorithm. Evaluation of the model was conducted using a 10-fold cross-validation process, where each time the model was trained using 90% of the data, and tested over the remaining 10%. The reported results are the mean of these 10 executions. Area Under the receiver operating characteristic Curve (AUC) was used as the main metric to assess the overall performance of the trained models.

## Data Availability

Access to the data used for this study can be made available upon request and is subject to internal review approval from the institutional review board of MHS with the current data sharing guidelines of MHS and Israeli law.

## Acknowledgments

The research was supported by the Israel Science Foundation (grant No. 3409/19) within the Israel Precision Medicine Partnership program, and the European Research Council (ERC) project #949850. The funders has no role in the design of the study, collection, analysis, and interpretation of data.

## Author information

### Contributions

ES and DY contributed to the study design. ES, RM, MP, TA, LY, MY and DY contributed in the analysis and interpretation of the results. TP, SG and SHG contributed in providing and interpreting the raw data. ES, MP, TA and LY wrote the code. ES, RM and DY wrote the first draft of the manuscript. All authors contributed to further versions of the manuscript. All authors have read and approved the manuscript.

