## Supplemental Material for "A Multilayer Model for Early Detection of COVID-19"

**Table S1. Description of the features considered in the machine learning models.**

| Layer | Feature | Description |
|---|---|---|
| **Layer 1: sociodemographic characteristics of the tested individual** | Age | A numeric feature indicating the age of the tested individual on January 1, 2020. |
| | Gender | A binary feature indicating the gender of the tested individual. |
| | Socioeconomic score | An ordinal feature that ranges between 1 and 10 and is defined by the Israeli Center Bureau of Statistics. The score per individual is determined based on the score of the statistical area of residence. Israel is stratified into ~3,500 statistical areas, corresponding to ~3000 residents per area. The statistical areas are homogenous in terms of their socioeconomic characteristics. |
| | Ethnicity | A categorical feature: 1) Ultra-orthodox Jews, 2) Arab individuals, and 3) the general population. The classification was determined based on the individual's self-reported information, the language used to receive healthcare services. If data was not available, ethnicity was determined based on the dominant ethnicity in the individual's residency area. |
| **Layer 2: spatiotemporal patterns of the disease** | Clinic-level positivity rate | A numeric feature indicating the positivity rate observed in the tested individual's clinic in the past 14 days (Maccabi has 137 clinics). |
| | Residency area-level positivity rate | A numeric feature indicating the positivity rate observed in the tested individual's residency area in the past 14 days. |
| | City-level positivity rate | A numeric feature indicating the positivity rate observed in the tested individual's city in the past 14 days. |
| | Socioeconomic-level positivity rate | A numeric feature indicating the positivity rate observed in the tested individual's socioeconomic level in the past 14 days. |
| **Layer 3: Individual medical history (EMRs, prior to the current COVID-19 test)** | Abnormal cardiovascular condition | A binary feature indicating whether the tested individual is diagnosed with an abnormal cardiovascular condition. |
| | Abnormal blood pressure | A binary feature indicating whether the tested individual is diagnosed with an abnormal blood pressure. The case definition of abnormal blood pressure condition as appeared in the electronic medical record is determined by one of the following:<br>A) Physician diagnosis.<br>B) Physician diagnosis and at least two blood pressure readings with values - systolic $\geq$ 140 and/or diastolic $\geq$ 90 prior or subsequent to the diagnosis.<br>C) At least 50% of the blood pressure readings values are - systolic $\geq$ 160 and/or diastolic $\geq$ 100.<br>D) Drugs consumption – at least six purchases of the following drugs: Thiazides, $\beta$-blockers, calcium channel blockers, and ACEs/ARBs. |
| | Cancer | A binary feature indicating whether the tested individual is diagnosed with cancer. |

| | | |
|---|---|---|
| | Diabetes | A binary feature indicating whether the tested individual is diagnosed with diabetes. |
| | Chronic kidney disease (CKD) | A binary feature indicating whether the tested individual is diagnosed with a chronic kidney disease. |
| | Chronic obstructive pulmonary disease (COPD) | A binary feature indicating whether the tested individual is diagnosed with a chronic obstructive pulmonary disease. |
| | Number of previous COVID-19 tests | The number of previous negative COVID-19 tests before the current one. |
| | Number of Diagnoses in the past five years | A numeric feature indicating the number of diagnoses in the past five years. |
| | Number of PCP visits in the past five years | A numeric feature indicating the number of PCP visits in the past five years. |
| | Number of hospitalizations in the past five years | A numeric feature indicating the number of hospitalizations in the past five years. |
| | Number of drug purchases in the past five years | A numeric feature indicating the number of drug purchases in the past five years. |
| | Number of vaccinations in the past five years | A numeric feature indicating the number of vaccinations in the past five years. |
| | Number of laboratory tests in the past five years | A numeric feature indicating the number of laboratory tests in the past five years. |
| | Number of months passed since the last diagnosis | A numeric feature indicating the number of months passed since the last diagnosis. If there were no diagnoses in the past five years, the value is set to None. |
| | Number of months passed since the last PCP visit | A numeric feature indicating the number of months passed since the last PCP visit. If there were no PCP visits in the past five years, the value is set to None. |
| | Number of months passed since the last hospitalization | A numeric feature indicating the number of months passed since the last hospitalization. If there were no hospitalizations in the past five years, the value is set to None. |
| | Number of months passed since the last drug purchase | A numeric feature indicating the number of months passed since the last drug purchases. If there were no drug purchases in the past five years, the value is set to None. |
| | Number of months passed since the last vaccination | A numeric feature indicating the number of months passed since the last vaccination. If there were no vaccinations in the past five years, the value is set to None. |
| | Number of months passed since the last laboratory test | A numeric feature indicating the number of months passed since the last laboratory test. If there were no laboratory tests in the past five years, the value is set to None. |
| | Number of previous COVID-19 tests | A numeric feature indicating the number of previous COVID-19 tests. |
| | BMI | The most recent BMI value recorded for the tested individual. |

| | | |
|---|---|---|
| | Referral for COVID-19 test | A binary feature indicating whether a referral for the COVID-19 test was issued. |
| **Layer 4: Information collected during the COVID-19** | Lives in the same household with an infected individual | A binary feature indicating whether the tested individual lives in the same household with an infected individual. |
| | Exposed to an infected individual | A binary feature indicating whether the tested individual was exposed to an infected individual (not necessarily in the same household) |
| | Home test | A binary feature indicating whether the test was performed at home |
| | Return from abroad | A binary feature indicating whether the tested individual returned from abroad |
| | Healthcare employee | A binary feature indicating whether the tested individual is an healthcare employee |
| | Identified via epidemiological investigation | A binary feature indicating whether the tested individual was identified following an epidemiological investigation of an infected individual. |
| | Fever > 37.5 | A binary feature indicating whether the tested individual reported on having a fever>37.5 symptom |
| | Cough | A binary feature indicating whether the tested individual reported on having a cough symptom |
| | Difficulty breathing | A binary feature indicating whether the tested individual reported on having a difficulty breathing symptom |
| | Muscles pain | A binary feature indicating whether the tested individual reported on having a muscles pain symptom |
| | Chest pain | A binary feature indicating whether the tested individual reported on having a chest pain symptom |
| | Extreme weakness | A binary feature indicating whether the tested individual reported on having an extreme weakness symptom |
| | Loss of taste and smell | A binary feature indicating whether the tested individual reported on having a loss of taste and smell symptom |
| | Diarrhea | A binary feature indicating whether the tested individual reported on having a diarrhea symptom |
| | Sore throat | A binary feature indicating whether the tested individual reported on having a sore throat symptom |
